# The German National Cohort: Ophthalmological Assessment, Baseline Profile and Potential for AI-based Eye Research

**DOI:** 10.64898/2026.05.04.26352019

**Authors:** Camila Roa, Ansgar Beuse, Alexandra Schweig, Sarah Müller, Klaus Berger, Caroline Brandl, Titus Brinker, Anne Elbrecht, Robert Finger, Gerd Geerling, Karin Halina Greiser, Carsten Grohmann, Kathrin Günther, Iris Heid, Andre Karch, Thomas Keil, Jessica Krepel, Michael Leitzmann, Claudia Meinke-Franze, Annette Peters, Sabine Schipf, Matthias Schulz, Alexander K. Schuster, Stefan N. Willich, Martin A. Leitritz, Marius Ueffing, Philipp Berens

## Abstract

**Objective:** To describe the ophthalmic examination protocol within the German National Cohort (NAKO) / NAKO Gesundheitsstudie, to report the baseline profile of participants undergoing ophthalmological assessment, and to illustrate the potential of these data as a population-based open resource for artificial intelligence (AI) research in eye health.

**Design:** Baseline analysis of ophthalmic data within the nationwide, population-based multicenter prospective NAKO study.

**Participants:** 48,460 adults in the ophthalmological level 2 module of 205,053 adults enrolled in NAKO, aged 19–74 years, with mean age 48.9 ± 12.5 years and 52.7% male.

**Methods:** All participants underwent standardized assessments of a wide range of biomedical examinations and detailed questionnaire-based data collection, including non-dilated color fundus imaging, visual acuity testing, recording of a brief ocular history. Ocular and systemic health measures were summarized using descriptive statistics. Fundus image quality and morphological features (e.g. cup-to-disc ratio, ateriole-to-venule-ratio) were assessed using open-source deep learning models. Standard deep learning architectures were trained on the fundus images to predict age, sex and blood pressure.

**Main Outcome Measures:** Percentage of fundus images graded as good quality; mean absolute error for age and blood pressure prediction; accuracy for sex prediction.

**Results:** The analysis includes 48,460 participants who successfully completed the level 2 ophthalmological baseline examination across 18 study sites in Germany. Mean visual acuity (logMAR) was 0.01 ± 0.20 (left eye) and 0.03 ± 0.21 (right eye). Self-reported ocular disease prevalence was 4.2% for cataract, 2.0% for glaucoma, and 0.9% for macular degeneration. 68.2% of fundus images were classified as gradable as a consensus of four deep learning-based quality grading models Morphological features such as cup-to-disc ratio and arteriole-to-venule-ratio showed systematic differences across age groups. Standard deep learning architectures showed comparative performance to the state-of-the-art for age, sex and blood pressure prediction (2.96 MAE for age prediction, 0.84 accuracy for sex prediction, 10.78 and 7.01 MAE for systolic and diastolic blood pressure prediction).

**Conclusions:** NAKO provides a large-scale, nationwide population-based resource with visual acuity measurements and systemic health indicators, as well as color fundus images in about 50,000 NAKO participants. The data sets the ground for studying eye health in the general adult population in Germany and can serve as a strong foundation for developing and validating AI tools in eye health research.

## Introduction

The German National Cohort (NAKO) / NAKO Gesundheitsstudie is a nationwide, population-based prospective study designed to investigate the determinants, risk factors, early markers, trajectories, and long-term consequences of major chronic diseases in Germany (Peters et al., 2022; Schipf et al., 2020). With 205,053 participants enrolled at baseline across 18 study centers, NAKO is one of the largest deeply phenotyped cohorts worldwide. It is among large population-based studies with over 100,000 participants, including nationwide studies such as the UK Biobank (Sudlow et al., 2015) and the China Kadoorie Biobank (Chen et al., 2011). NAKO’s comprehensive design incorporates multimodal imaging, biospecimen collection, environmental and behavioural profiling, and a planned overall duration of 25 – 30 years (Peters et al., 2022; The National German Cohort Consortium, 2014). A subgroup of more than 55,000 participants was offered an extended examination programme (so called level 2 examinations). Within this framework, the ophthalmology module was established as one of the level 2 investigation with three key objectives:

i. to characterize visual acuity and self-reported ocular diseases.
ii. to characterize the prevalence and incidence of ophthalmic findings on imaging;
iii. to explore the associations between ocular measures and systemic health indicators, as well as the reverse.

In this paper, we describe the ophthalmic examinations in the NAKO and outline the standardized procedures used to collect eye-related data. We analysed baseline non-imaging findings and self-reported eye health characteristics and– using open-source deep learning models -evaluated the quality of the acquired fundus images and extracted key morphological features. Additionally, we trained standard deep learning models to predict age, sex and blood pressure to demonstrate the suitability of the fundus images taken as part of NAKO for AI-based eye health and oculomics research.

## Methods

### Design, Baseline Recruitment and Ethical Approval

The NAKO is a large, prospective, population-based cohort study conducted in Germany. Participants were randomly selected from compulsory population registries in 16 German regions using age- and sex-stratified sampling. They were examined between 2014 and 2019 in 18 study centers representing urban, suburban, and rural settings (Rach et al., 2025). The NAKO aimed to recruit approximately 200,000 residents between the ages of 20 and 69 (The National German Cohort Consortium, 2014).

The baseline recruitment yielded 205,415 participants aged 19–74 years. Inclusion criteria were: residing in the study catchment area, being within the target age range at the time of invitation, ability to provide written informed consent, and proficiency in the German language to understand questionnaires and instructions. Exclusion criteria included being unable to consent. The overall response rate averaged 15.6%, with higher participation among older individuals and women with differences across regions (Peters et al., 2022; Rach et al., 2025). Recruitment was monitored to ensure representativeness across predefined strata. After 362 participants withdrew informed consent, the sample consisted of 205,053 individuals as of October 2019 (Peters et al., 2022).

Baseline assessment included standardized, computer-assisted face-to-face interviews, self-administered questionnaires, standardized physical and medical examinations, and biosample collection. A core set of questionnaires and examinations was offered to all participants (so called level 1 modules), and an extended set of in-depth examinations and questionnaire modules (level 2 modules) was offered to a 20% random sample of all participants plus to additional participants undergoing magnetic resonance imaging at 5 of the 18 study centres. For training of the AI-based models regarding estimation of age, sex and blood pressure, we used data obtained during interview and during measurement of the sitting blood pressure with the OMRON-HEM 705 IT (Peters et al, 2022).

The study protocol of NAKO was approved by the ethics committees of all participating institutions, and all participants provided written informed consent in accordance with German legal and data protection requirements. It is conducted in accordance with the Declaration of Helsinki and national standards for good clinical and epidemiological practice (Peters et al., 2022).

### The Ophthalmology Module

The intensified level 2 examination of NAKO includes an ophthalmology module where participants received a eye assessment of visual acuity testing and color fundus imaging. It was conducted in 56,971 participants of the baseline examination (**Figure 1**). Of those participants, 48,460 completed at least with one eye an visual acuity and and retina photography, as mobilized by the Transfer Hub (**Figure 1**, **Table 1**).

**Figure 1:**
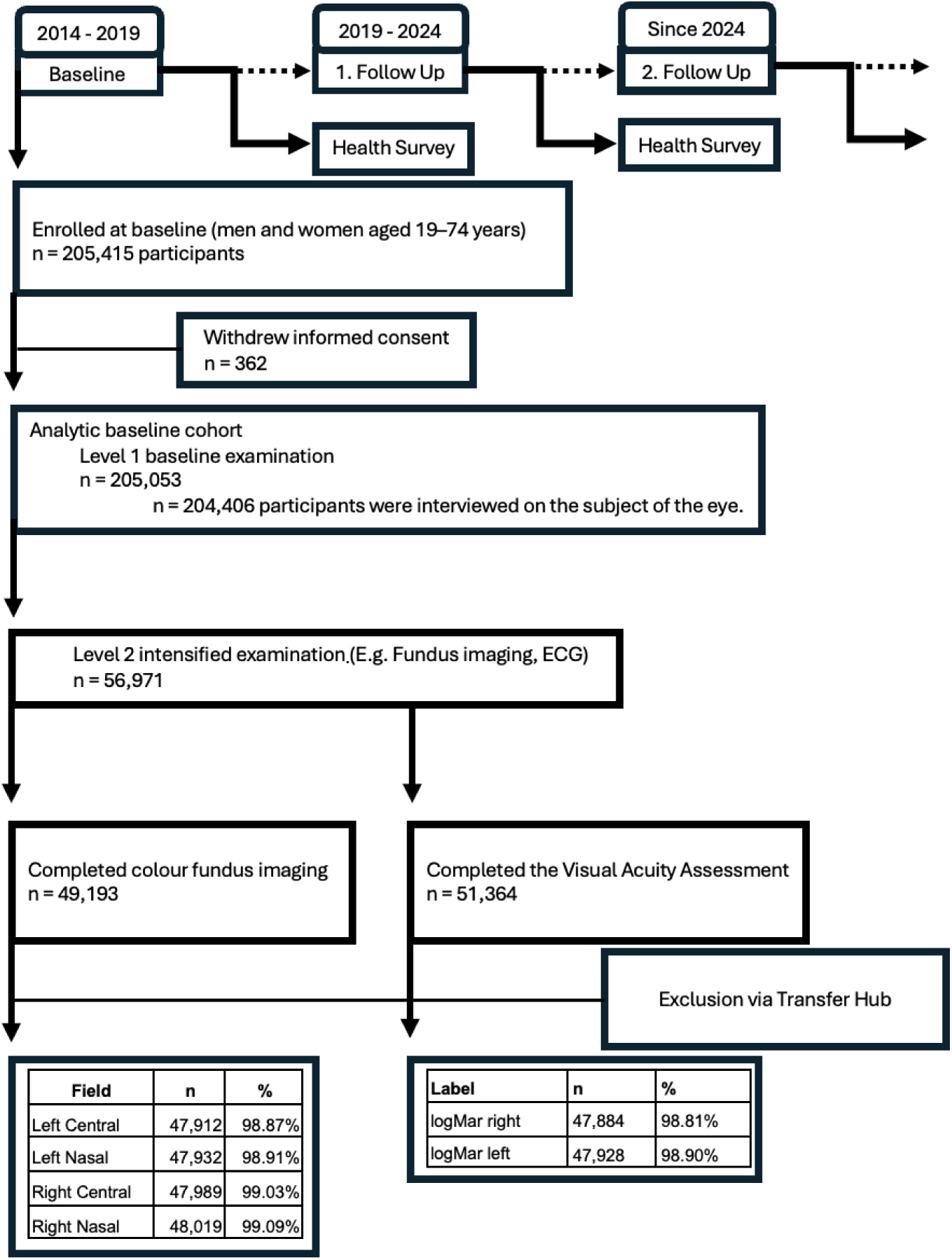
Flowchart of study participants for the basline examination in the Level 2 ophthalmology submodule of the German National Cohort (NAKO). Numbers are reported as in n = participants

**Table 1:** Summary of demographic, systemic and eye health variables of n=48,460 participants mobilized by NAKO TransferHub from the baseline in the NAKO Ophthalmology dataset. Continuous variables are reported as mean ± sd and discrete variables as n (%). Note: Percentages are relative to the total number of participants.

| Label | Mean | STD | Min | Max |
| --- | --- | --- | --- | --- |
| Age (years) | 48.91 | 12.53 | 19 | 74 |
| Systolic Blood Pressure (mmHg) | 127.14 | 16.16 | 75 | 236 |
| Diastolic Blood Pressure (mmHg) | 78.39 | 9.96 | 44 | 136 |
| Left eye logMAR | 0.01 | 0.20 | -0.20 | 1.20 |
| Right eye logMAR | 0.03 | 0.21 | -0.20 | 1.20 |
| Sex | Male: 52.69%, Female: 47.31% |  |  |  |
| Smoking status | Not a smoker: 47.12%, Former smoker: 31.58%, Smoker: 18.95%, Missing: 2.34% |  |  |  |
| High Blood Pressure | No: 76.05%, Yes: 23.87%, Missing: 0.09% |  |  |  |
| Diabetes Mellitus | No: 94.96%, Yes: 4.91%, Missing: 0.13% |  |  |  |
| Cataracts | No: 95.54%, Yes: 4.21%, Missing: 0.25% |  |  |  |
| Glaucoma | No: 97.68%, Yes: 2.00%, Missing: 0.31% |  |  |  |
| Macular Degeneration | No: 98.78%, Yes: 0.89%, Missing: 0.33% |  |  |  |
| Site | Augsburg: 17.94%, Neubrandenburg: 11.91%, Mannheim: 10.09%, Essen: 9.02%, Bremen: 4.27%, Leipzig: 3.99%, Halle: 3.97%, Münster: 3.96%, Freiburg: 3.95%, Berlin Mitte: 3.77%, Berlin Süd: 3.76%, Berlin Nord: 3.34%, Dusseldorf: 3.32%, Saarbrücken: 3.27%, Regensburg: 3.24%, Hannover: 3.21%, Kiel: 2.55%, Hamburg: 2.50%, Neustrelitz: 0.89%, Demmin: 0.86%, Waren (Müritz): 0.18% |  |  |  |

Within NAKO, a panel of ophthalmic, epidemiological and information technological experts from Germany (NAKO eye expert group) is responsible for the development of the ophthalmology program, oversight, and support of future project development. For the development of the ophthalmology program, a pre-test study was conducted (Leitritz et al., 2013b), testing standard operating procedures (SOPs) for visual acuity assessment and color fundus imaging as well as different fundus cameras. SOPs were modified to maximize information content versus participants’ time in study center to ensure feasibility at the scale of NAKO.

The NAKO-retina competence unit is responsible for overall management and quality assurance of NAKO eye data. This includes expert oversight of conduct, quality management and reporting to participants across 18 study centers, and assessment of incidental findings and quality management of eye examinations among all 18 study centers, ensuring continuous quality assessment through ongoing communication with the central data collection units, quality management and study centers. It is tasked with overseeing the overall management and quality assurance of the eye data. This includes conducting regular (remote) site visits ensuring appropriate and equal on-site conditions and training sessions to ensure the integrity and accuracy of both examinations and data processing.

All examination procedures followed SOPs and all study personnel were trained and certified before collecting data from participants. Visual acuity was tested monocularly while participants wore their walk-in distance correction if needed (glasses or contact lenses). In addition, participants held a stenopaic hole in front of the tested eye, which contained a central aperture and an additional rotatable insert with several 2.0 mm perforated openings while the non-tested eye was occluded. This facilitates further letter recognition and lowers the impact of residual refractive errors. Illuminated optotype charts with Latin letters (ETDRS Visual Acuity Tester Table A, Steinbeis Transfer Center for Biomedical Optics, Tübingen, Germany), based on the ETDRS protocol, were used. The testing distance was 4 m, allowing assessment of visual acuity from 0.05 to 1.0. In case of examination room length restrictions, a fixed mirror was used to create an overall reading distance of 4 m between participant and monitor. The test was terminated when less than three of five letters were correctly identified. Results (type of refractive correction, identified optotypes, and notable observations) were digitally documented with responses marked by study staff (Leitritz et al., 2013a). The visual acuity test was harmonized with and adapted from the UK biobank examination protocol to permit future joint analyses.

Retinal imaging was performed using a fundus camera (model DRS s/n 7170, Revenio group, icare: icare-world.com, Finland, formerly CenterVue S.p.A.) without mydriasis. The device supports single- and multi-field imaging with automated alignment. Images were captured with a field of view of 45° x 40°. The camera has an autofocus range of −15 to +15 D, internal fixation LEDs, a working distance of 37 mm, and a 5-megapixel sensor (2592 x 1944 pixels), yielding a resolution of 48 pixels/degree and a retinal resolution of 15 µm² for the standard eye.

For each eye, one macula-centered image and one optic disc–centered image was usually acquired. In case of obviously insufficient image quality, the study nurses repeated the respective image acquisition, and all images were saved. The imaging procedure required no medication-induced dilation of the pupil (mydriasis), was painless and caused only minor inconvenience (brief glare) for the participants. Fundus images were acquired in a darkened room by trained and regularly recertified study staff, according to a standardized SOP.

Following approval for pseudonymized analysis, the images were transferred to the Reading Center at the Department of Ophthalmology, University Hospital Tübingen, Germany. Here, manual grading and annotation of the images from a list of predefined findings was performed. These annotations are being made available through the NAKO Transferhub. The ophthalmologic reading centre in Tübingen (Germany) provides specialized personnel, state-of-the-art equipment, and expert knowledge to support the computer-assisted, standardized evaluation of data. Its organisational framework facilitates image analysis, which is enhanced by advanced computational image analysis tools.

Exclusion criteria for visual acuity testing and fundus imaging were assessed prior to testing and included participant refusal, severe ocular infection, ocular surgery within prior four weeks, prosthetic eye, known blindness, and (for fundus imaging) photosensitive epilepsy. Testing could be discontinued at any time by the participant, examiner, or due to technical issues (e.g., camera malfunction). The complete eye examination (visual acuity and fundus imaging) required approximately 10 minutes.

Alongside the eye examination, participants were assessed for self-reported macular degeneration, cataract, optic nerve inflammation, and glaucoma using a standardized questionnaire.

All data is accessible through the official NAKO Transfer Hub (https://transfer.nako.de), enabling broad scientific use under established governance and data-protection frameworks. The study structure also allows the integration of ancillary sub-cohorts to address emerging research questions when needed.

### Statistical Analysis

We performed a descriptive analysis of the study population, summarizing basic demographic information (sex, age, study site), general health related variables (blood pressure, smoking status, diabetes) and ophthalmological variables (visual acuity, presence of eye diseases in self-report). Continuous variables are presented as means and standard deviations (SD) as well as minimum and maximum values. Categorical variables were summarized using absolute counts and percentages. Age-group distributions were calculated separately for men and women and reported as proportions of the total sample. Missing data were reported explicitly and no imputation was performed. All analysis were performed in Python using standard statistical procedures.

### Automated Fundus Image Analysis

#### Preprocessing

We used the Fundus Image Toolbox (Gervelmeyer et al., 2025) to crop and center the fundus images appropriately.

#### Automatic Assessment of Image Quality

To automatically assess the quality of the fundus images, we used four different open-source software packages that include quality prediction models. For all models, we used default parameters and applied them to the pre-processed fundus images resized to 512 x 512.

- Fundus Image Toolbox (Gervelmeyer et al., 2025): a python package for fundus image processing that includes quality grading, fovea and optic disc localization, vessel segmentation, registration and circle cropping. Quality is rated on a scale from 0 to 1 (ungradable to gradable) by an ensemble of standard deep learning architectures (ResNets and EfficientNets) trained on the DeepDRiD and DrimDB datasets (R. Liu et al., 2022; Oraá et al., 2020). We used the default threshold of 0.25, classifying images with higher scores as gradable.
- AutoMorph (Zhou et al., 2022): a pipeline for automated analysis of retinal images that includes preprocessing, quality grading, vessel, artery, vein, and disc segmentation and morphological feature measurement. For quality grading, a standard deep learning architecture (EfficientNet-B4) trained on the EyeQ dataset (Fu et al., 2019) classifies images as good, usable, or reject. We considered good and usable images as gradable.
- VascX (Vargas Quiros et al., 2025): a pipeline for fundus analysis that includes preprocessing, quality grading, vessel artery-vein and disc segmentation, and fovea localization. The quality grading model is an ensemble of fine-tuned deep learning models (ResNet101) on the EyeQ dataset that classifies images as good, usable or reject. We considered good and usable images as gradable.
- RetiRatey (https://github.com/justinengelmann/RetiRatey): a fine-tuned small convolutional variant of the MobileNet-V4 family that rates the quality of retinal images on a scale from 1 to 4 (1: Very good, 2: Good, 3: Borderline 4: Bad) for different regions of interest: the macula, the disc, and the vessels. We considered images with scores lower than 3 on all three regions of interest as gradable.

Automatic gradings by all four methods will be made available via the NAKO Transfer Hub.

#### Automatic Extraction of Morphological Fundus Image Features and Epidemiological Analysis

We used the AutoMorph toolbox (Zhou et al., 2022) to extract morphological measurements from the macula-centred retinal images. We focussed on the following measurements: the vertical cup to disk ratio, along with the Central Retinal Artery Equivalent (CRAE), the Arteriolar to Venular Ratio (AVR), and the vessel density of zone b (Knudtson et al., 2003). For these variables, we analyzed their relationship with age and sex using generalized additive models (GAMs) via the R package mcgv (version 1.9-4), fitting a smooth term for age, which was allowed to differ between sexes, resulting in the formulamodel `y ∼ sex + s(age, by=sex)`, where y is the variable of interest (Wood, 2017). We removed outliers using the interquartile range (IQR) method, applied independently to each feature. Samples below Q1 − 1.5×IQR or above Q3 + 1.5×IQR were excluded, where IQR=Q3 -Q1. Extracted morphological properties will be made available through the NAKO Transfer Hub.

#### Deep Learning-based Prediction of Systemic Variables

We trained several standard deep learning models – ResNet18, ResNet50, Inceptionv3, Xception, EfficientNetv2-Small, ViT-Small and Swin-Tiny (Chollet, 2017; Dosovitskiy et al., 2020; He et al., 2016; Z. Liu et al., 2021; Szegedy et al., 2016; Tan & Le, 2021) – to predict age, blood pressure and sex from fundus images from the NAKO baseline examination. The size of the transformer models was chosen to yield models of a size in the same order of magnitude as a ResNet50. Preliminary experiments revealed that ImageNet pretraining was helpful to obtain better performing models in less time. Thus, all models were initialized with ImageNet weights and fine-tuned for a maximum of 25 epochs with early stopping. We used a batch size of 256 and an AdamW optimizer with a learning rate of 10^−4^ and weight decay of 10^−4^. We trained the classifiers with cross-entropy loss and the regression models with mean squared error loss (MSE). The blood pressure model was trained with two prediction heads as a multi-task model for both systolic and diastolic blood pressure. All trained models will be made available through the NAKO Transfer Hub.

For development of deep learning models, we used images that were graded as “good” by the Fundus Image Toolbox (Gervelmeyer et al., 2025). We always selected the highest-quality image available for each participant and eye. This resulted in a total of 153,134 images from 45,247 participants. The images were resized to 224 x 224 pixels and split into training set (90%) and test set (10%). All models were trained using 5-fold cross-validation on the training split, with one of the folds serving as the validation set. We report the average performance and the 95% confidence intervals (CIs), computed as the 2.5 and 97.5 percentiles for various metrics, including accuracy and area under the receiver operating characteristic curve (AUC) for sex and mean absolute error (MAE) and the coefficient of determination (R^2^) for age, systolic blood pressure, and diastolic blood pressure. We selected best model based on validation performance metrics and then evaluated it on the test set using a bootstrap procedure with 1,000 resamples of 15,000 images from the test set.

## Results

### Population Profile

Within the data mobilized by the NAKO Transfer Hub 48,460 participants took part in the baseline ophthalmology module and had at least one fundus image for at least one eye and visual acuity examination taken (**Figure 1**). Of these, 25,534 were male men (52.7%) and 22,926 were female women (47.3%) (**Table 1**). Thus, the percentage of women receiving the baseline eye exam was lower than the percentage of women in the overall NAKO study (50.5%) (Stein et al., 2024). The mean age of the participants was 48.9 ± 12.5 years, ranging from 19 to 74 years old, similar to the overall NAKO average (49.9 ± 12.8 years). The different age ranges were well represented: 21.6% of participants were between 20 and 39 years old; 27.8% of participants were between 40 and 49 years old; 27.4% were between 50 and 59 years old; 21.5% were between 60 and 69 years old; and 1.7% of participants were between 70 and 79 years old (Supplementary Table 1). This made the cohort comparatively young at the start of the study (Peters et al., 2022; Sudlow et al., 2015). Participants from all study centers completed the ophthalmological assessment (**Table 1**).

The cardiovascular and metabolic profile of the participants was typical of a comparatively young adult population cohort (see **Table 1**; Supplementary Tables 2-3): The mean systolic blood pressure was 127.1 ± 16.2 mmHg (mean ± SD, range 75–236) and the mean diastolic blood pressure was 78.4 ± 10.0 mmHg (range 44–136), with slightly higher systolic blood pressure in men than in women. Based on self-reports, 23.9% of participants reported high blood pressure, while 76.0% reported no history of hypertension. Additionally, 19.4% reported to be active smokers, and 32.6% had smoked in the past, with fewer past smokers in women than in men. In total, 4.9% of participants reported to have Diabetes mellitus, with less women than men.

The participants reported a variety of ocular diseases, including cataracts (4.2%), glaucoma (2.0%), and macular degeneration (0.9%) **(Table 1)**. Visual acuity (, assessed using logMAR) measurements, was on average 0.01 ± 0.20 in the left eye on average (mean ± SD) and 0.03 ± 0.21 in the right eye, with ranges from −0.20 to 1.20 in both eyes, and similar values for men and women. Of the 48,460 participants, 203,493 fundus images were taken, 51,140 from the left central field, 51,112 from the left nasal field, 50,348 from the right central field, and 50,893 from the right nasal field. As per instruction in the SOPs, images were re-taken 1 to 2 times for some participants, if the first image was judged to be of insufficient quality; we included all of these images in the dataset.

### Quality of Fundus Images

We found that the acquired color fundus images were generally of high quality. However, there were also some images of insufficient quality for grading (**Figure 2**). To systematically evaluate the quality of the acquired fundus images and its dependency on age, we automatically analysed all 203,493 fundus images of the 48,460 participants with four deep learning models for fundus image quality assessment (see Methods). Overall, the four algorithms FIT, RetiRatey, AutoMorph and VascX graded 155,857 (76.6%), 177,172 (87.1%), 154,542 (75.9%) and 151,415 (74.4%) out of 203,493 images as gradable, and for 138,885 out of 203,493 (68.2%) all four algorithms agreed that these images were of high quality. This percentage is higher than that of other population-based cohort studies, such as the ophthalmological cohort of UK Biobank (high quality in agreement of all four algorithms: 48.7%) (Chua et al., 2019) assessed by us using the same algorithms and settings. Depending on the algorithm and the fundus image field, we found that between 70.1% and 91.4% of participants had at least one image for at least one eye that was classified as good quality (**Table 2**). Interestingly, the proportion of gradable images was typically higher for the central fields in both eyes, and image quality correlated with with the order in which images were taken (right central, right nasal, left central, left nasal). Image quality declined with increasing age (from 80-90% gradable images below 40 years to 40.8% at oldest age-group; Supplementary Table 4) similarly in men and women.

**Figure 2:**
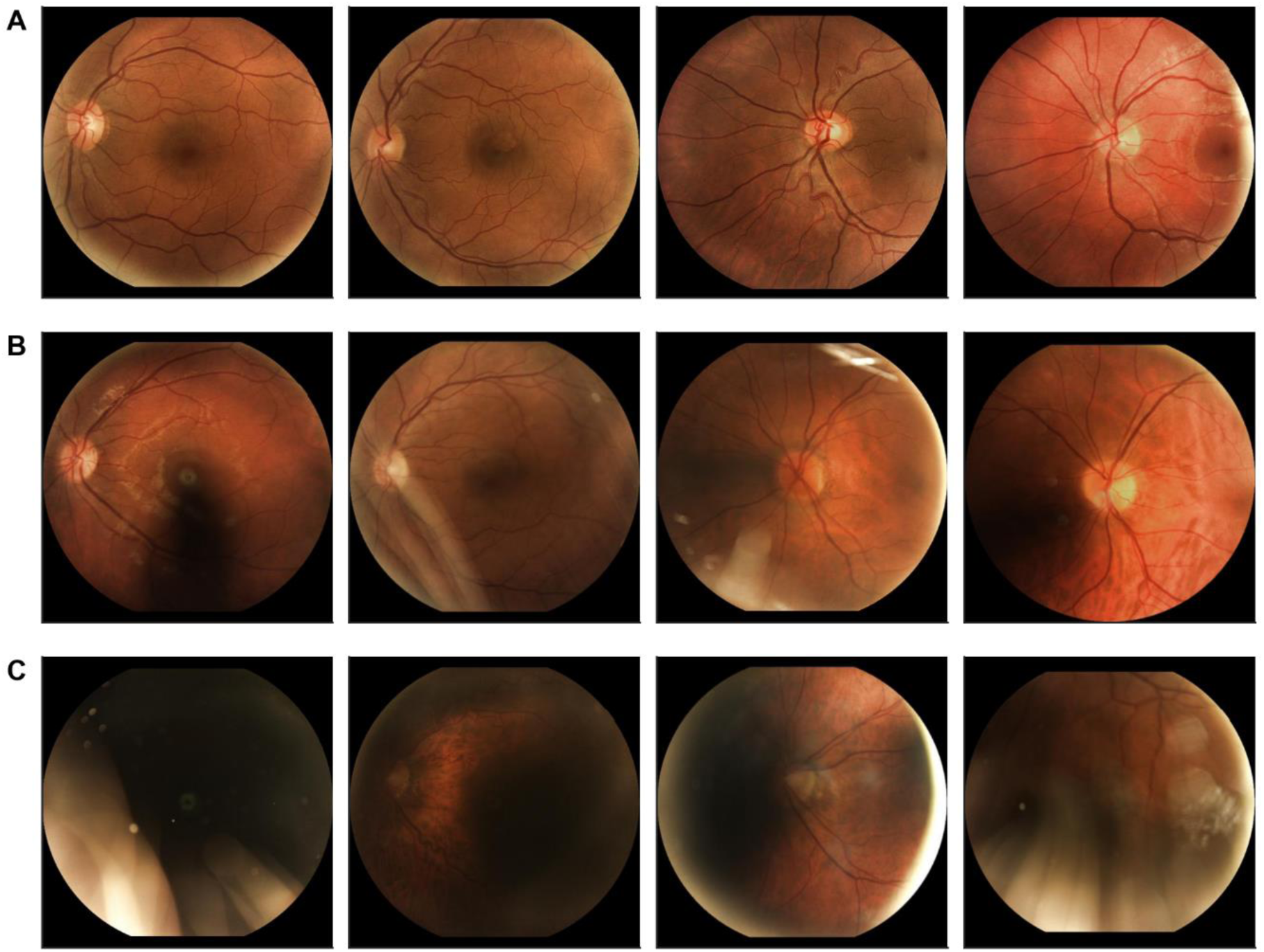
Example Images from the NAKO dataset. A. Fundus images with good quality, as judged by automatic quality grading by the Fundus Image Toolbox algorithm. From left to right: Female, in their 40s; Male, in their 40s; Male, in their 30s; Male, in their 20s. (as reported during the examination). B. Fundus images with borderline quality, showing evidence of artefacts or poor illumination. From left to right: Female, in their 30s; Female, in their 40s; Female, in their 70s; Male, in their 40s. C. Fundus images with insufficient quality not used for further analysis. From left to right: Male, in their 50s; Male, in their 60s; Male, in their 60s; Male, in their 50s.

**Table 2:** Percentage of participants (n=48,460) with gradable images according to four different AI models for quality grading (FIT, RetiRatey, AutoMorph, VascX) and consensus rating for participants with gradable images according to all models (All).

| Field | FIT | RetiRatey | AutoMorph | VascX | All |
| --- | --- | --- | --- | --- | --- |
| Right Central | 83.1 | 90.4 | 85.4 | 87.7 | 81.3 |
| Right Nasal | 83.6 | 91.4 | 80.3 | 72.3 | 68.2 |
| Left Central | 76.7 | 84.7 | 77.9 | 82.3 | 74.9 |
| Left Nasal | 72.6 | 89.2 | 70.1 | 64.9 | 58.5 |

### Epidemiological Analysis of Basic Morphological Fundus Properties

To demonstrate the potential of the dataset for epidemiological analysis, we used an automatic, externally validated deep learning tool (Zhou et al., 2022) to extract standardized morphological measurements related to the optic nerve and microvascular health of the fundus, such as the vertical cup-to-disc ratio, central retinal artery equivalent (CRAE), arteriole-to-venule ratio (AVR) and vessel density (see Methods). On average, vertical cup-to-disk ratio was 0.501, CRAE was 154 μm, AVR was 0.679 and vessel density was 0.00689. We examined age- and sex-related differences in retinal and optic nerve head measures. Cup-to-disc ratio increased with age, whereas CRAE, AVR, and vessel density declined from early adulthood (Figure 3). Sex differences were statistically significant but small in magnitude: women showed slightly higher AVR and vessel density and lower cup-to-disc ratios at younger ages, with age-related differences largely similar between sexes and convergence most evident for cup-to-disc ratio at older ages (see Supplementary Tables 5-8).

**Figure 3:**
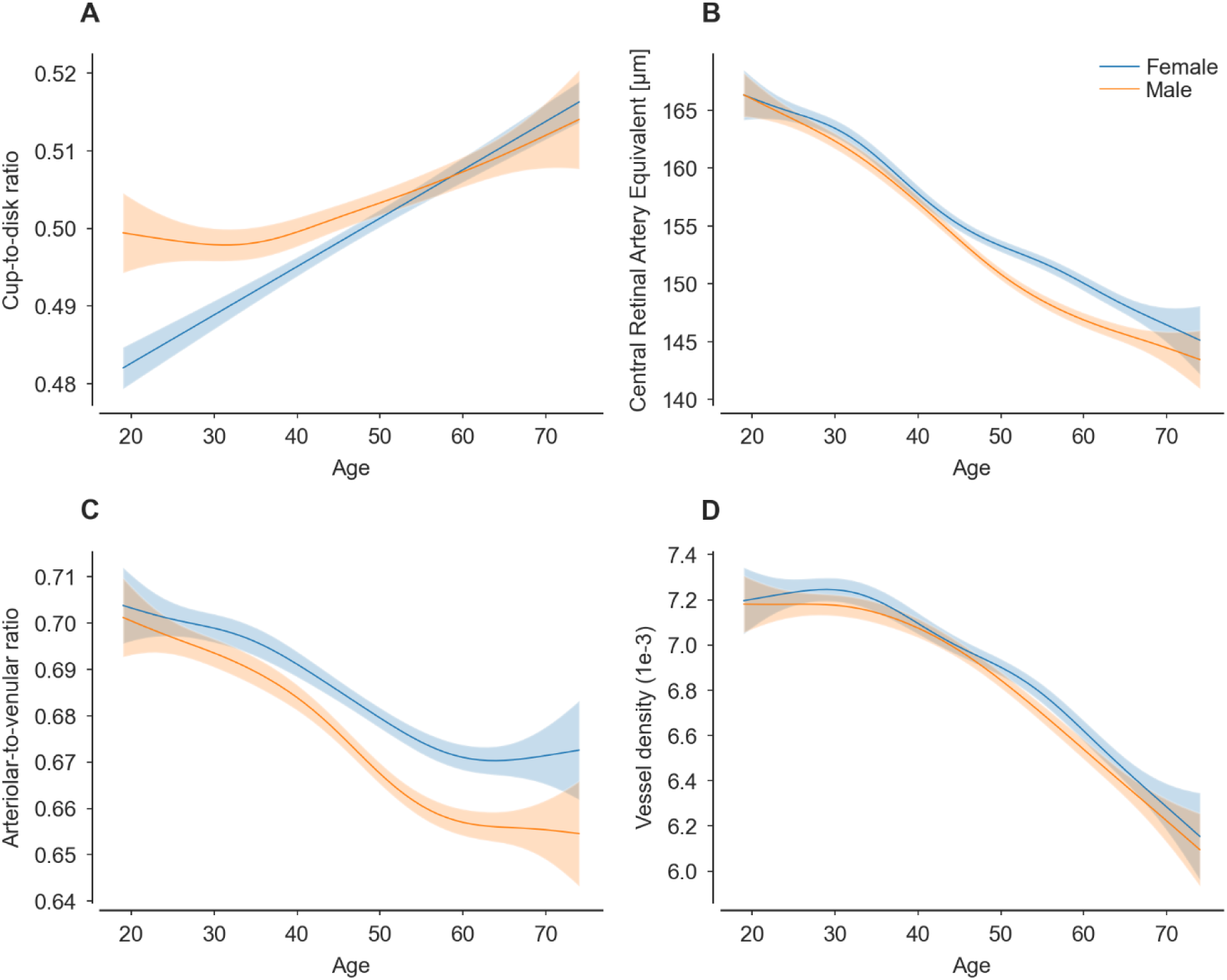
Morphological features extracted from fundus images show dependence on life course and sex. A. Cup-to-disc ratio. Solid lines show mean prediction of a fitted generalized additive model (GAM), with standard errors. n= 28,220. B. CRAE Knudson. n= 27,297. C. Arterial-to-vein ratio Knudtson (AVR). n= 27,114. D. Vessel density. n= 28,608. Detailed statistical information about the fitted GAM models is available in the Supplementary Material.

### Deep Learning Models for Age, Sex and Blood Pressure Prediction

To demonstrate the dataset’s potential for AI-based ophthalmology and oculomics research, we trained various deep learning models including convolutional and transformer architectures to predict age, sex and blood pressure from the fundus images based on the data recorded during the baseline examination. These models included ResNet18, ResNet50, Inceptionv3, Xception, EfficientNetv2-Small, ViT-Small and Swin-Tiny. We found that all architectures could predict these variables with similar or better accuracy than reported in the literature (Poplin et al., 2018) (**Table 3** and **Table 4**). For instance, age prediction accuracy ranged from 2.964 years (2.920-3.005 95% CI; mean absolute error [MAE]) for the EfficientNet to 3.181 MAE (3.138 – 3.223) for the Xception network (**Figure 4**). For systolic blood pressure prediction, performance ranged from 10.781 mmHg MAE (10.644 -10.922, 95% CI) for the EfficientNet to 11.419 mmHg MAE (11.274 – 11.585, 95% CI) for the Swin transformer, indicating substantial unexplained variability. The EffificentNetv2 network and the ResNet50 typically performed best (**Table 3** and **Table 4**), making them good starting points for oculomics approaches.

**Figure 4:**
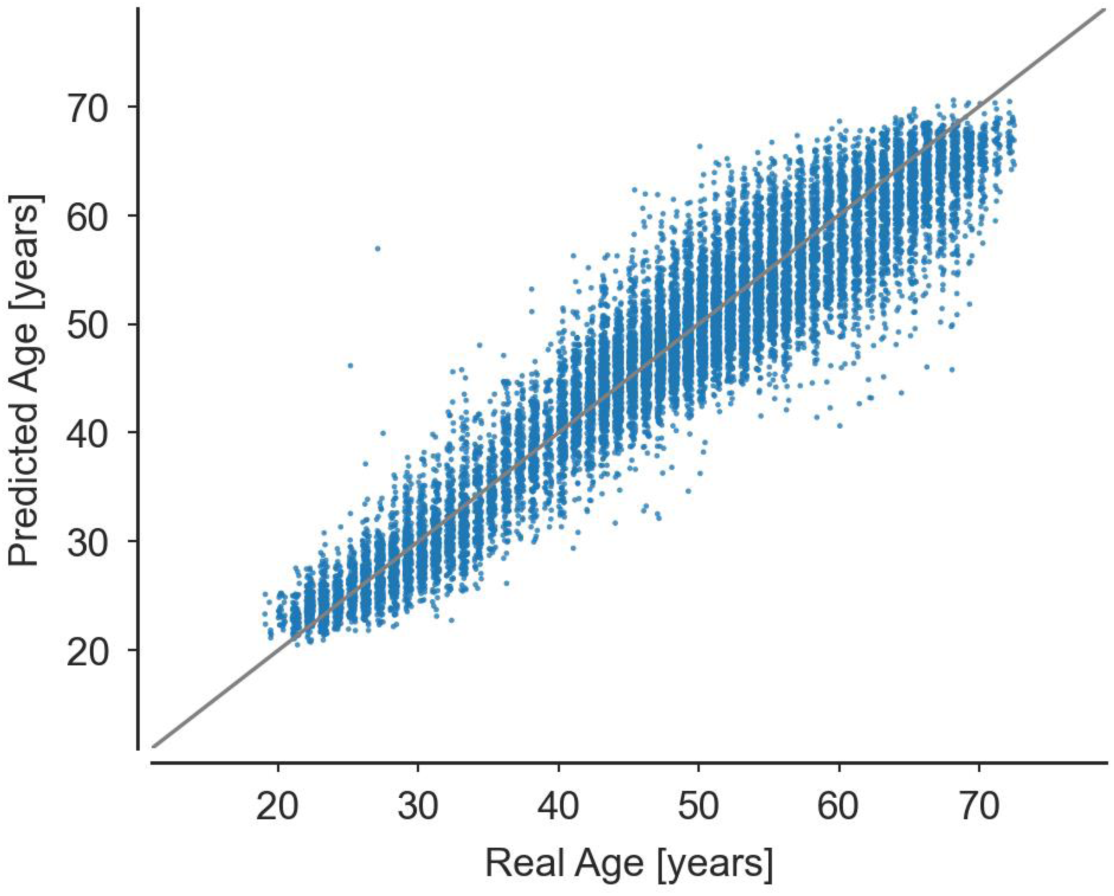
Real vs. predicted age for n=15,245 NAKO participants for an EfficientNetv2-S model. r²=0.901, MAE=2.965.

**Table 3:** Performance of different deep learning architectures for prediction of continuous variables (age, systolic blood pressure, diastolic blood pressure), measured by the mean absolute error (MAE) and r², with 95% confidence intervals. For comparison, literature values from Poplin et al. trained on UKB and EyePACS and evaluated on a hold-out set from UKB.

| Target | Model | MAE | R2 |
| --- | --- | --- | --- |
| Age | Resnet18 | 3.104 (3.063 - 3.148) | 0.889 (0.885 - 0.893) |
|  | Resnet50 | 3.079 (3.037 - 3.121) | 0.893 (0.889 - 0.896) |
|  | Inceptionv3 | 3.113 (3.074 - 3.154) | 0.889 (0.885 - 0.893) |
|  | Xception | 3.181 (3.138 - 3.223) | 0.885 (0.881 - 0.889) |
|  | EfficientNetv2-S | 2.964 (2.920 - 3.005) | 0.901 (0.898 - 0.904) |
|  | ViT-Small | 3.101 (3.057 - 3.144) | 0.890 (0.886 - 0.894) |
|  | Swin-Tiny | 3.079 (3.038 - 3.121) | 0.894 (0.891 - 0.898) |
|  | Poplin et al (Inceptionv3) | 3.26 (3.22,3.31) | 0.74 (0.73,0.75) |
| Systolic blood pressure | Resnet18 | 10.923 (10.789 - 11.052) | 0.210 (0.196 - 0.223) |
|  | Resnet50 | 10.853 (10.711 - 10.995) | 0.210 (0.196 - 0.224) |
|  | Inceptionv3 | 10.950 (10.807 - 11.101) | 0.199 (0.186 - 0.213) |
|  | Xception | 10.906 (10.764 - 11.049) | 0.188 (0.174 - 0.202) |
|  | EfficientNetv2-S | 10.781 (10.644 - 10.922) | 0.215 (0.201 - 0.229) |
|  | ViT-Small | 10.970 (10.829 - 11.109) | 0.200 (0.187 - 0.212) |
|  | Swin-Tiny | 11.419 (11.274 - 11.585) | 0.097 (0.083 - 0.111) |
|  | Poplin et al (Inceptionv3) | 11.35 (11.18,11.51) | 0.36 (0.35,0.37) |
| Diastolic blood pressure | Resnet18 | 6.969 (6.890 - 7.051) | 0.204 (0.190 - 0.218) |
|  | Resnet50 | 6.970 (6.885 - 7.051) | 0.205 (0.192 - 0.217) |
|  | Inceptionv3 | 7.090 (7.006 - 7.171) | 0.180 (0.167 - 0.193) |
|  | Xception | 7.067 (6.976 - 7.152) | 0.186 (0.171 - 0.199) |
|  | EfficientNetv2-S | 7.014 (6.931 - 7.099) | 0.201 (0.187 - 0.214) |
|  | ViT-Small | 6.967 (6.888 - 7.049) | 0.204 (0.191 - 0.217) |
|  | Swin-Tiny | 7.242 (7.155 - 7.327) | 0.130 (0.117 - 0.143) |
|  | Poplin et al (Inceptionv3) | 6.42 (6.33,6.52) | 0.32 (0.30,0.33) |

**Table 4:**
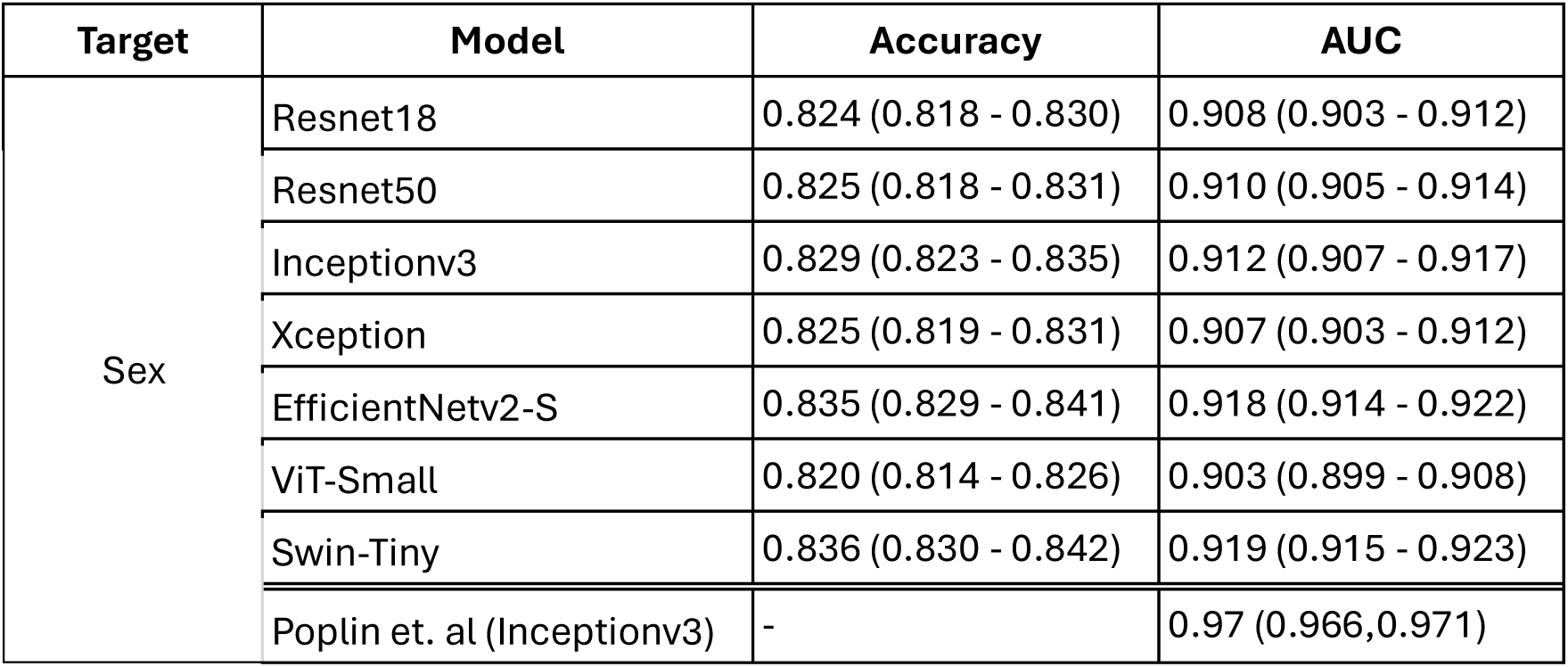
Performance of different deep learning architectures for prediction of the binary variable sex, measured by the accuracy and AUC, with 95% confidence intervals. For comparison, literature values from Poplin et al. trained on UKB and EyePACS and evaluated on a hold-out set from UKB.

| Target | Model | Accuracy | AUC |
| --- | --- | --- | --- |
| Sex | Resnet18 | 0.824 (0.818 - 0.830) | 0.908 (0.903 - 0.912) |
|  | Resnet50 | 0.825 (0.818 - 0.831) | 0.910 (0.905 - 0.914) |
|  | Inceptionv3 | 0.829 (0.823 - 0.835) | 0.912 (0.907 - 0.917) |
|  | Xception | 0.825 (0.819 - 0.831) | 0.907 (0.903 - 0.912) |
|  | EfficientNetv2-S | 0.835 (0.829 - 0.841) | 0.918 (0.914 - 0.922) |
|  | ViT-Small | 0.820 (0.814 - 0.826) | 0.903 (0.899 - 0.908) |
|  | Swin-Tiny | 0.836 (0.830 - 0.842) | 0.919 (0.915 - 0.923) |
|  | Poplin et. al (Inceptionv3) | - | 0.97 (0.966,0.971) |

## Discussion

We characterized the design and methodological framework of the ophthalmology module of the German National Cohort (NAKO) (The National German Cohort Consortium, 2014) and present baseline results from 48,460 participants who underwent standardized visual acuity testing and nonmydriatic color fundus imaging, as well as extensive biomedical examination. With its scale, NAKO is among major international population-based cohorts in ophthalmology such as the UK Biobank baseline ophthalmology subset with around 68.000 participants with OCT and color fundus maging (Warwick et al., 2023). Within Europe, the study will rank among the largest deeply phenotyped population-based nationwide cohorts, complementing more local, well-established cohorts in Germany with extensive phenotyping and ophthalmological assessments, such as the Gutenberg Health Study (GHS) (Höhn et al., 2015; Wild et al., 2012), the AugUR study (Brandl et al., 2019; Stark et al., 2015), the Hamburg City Health Study (HCHS) (Beuse et al., 2025; Jagodzinski et al., 2020), the Life Adult Study (Loeffler et al., 2015), the Rhineland Study (Mauschitz et al., 2022) and the KORA study (Holle et al., 2005). In addition to the level 2 ophthalmology module, the NAKO includes local in-depth level 3 modules with ophthalmological relevance (such as for dry eye disease) in selected study centers.

The mean logMAR values, which were close to 0.0, indicate that this community-based sample predominantly has normal vision. These values are broadly consistent with values reported in large European population-based cohorts, particularly among individuals without significant ocular pathology (Chua et al., 2019; Höhn et al., 2015). These visual acuity assessments in NAKO will become particularly important when follow-up investigations will be able to compare changes over time as individuals age and derive determinants of this aging. Similarly, color fundus images at baseline jointly with follow-up imaging will also be key to derive determinants of eye health changes as well as ocular presentations of other diseases. Together with objectively measured eye parameters, self-reported eye disease can provide insights into the awareness of eye diseases in the general population.

Self-reported eye disease can provide insight into disease awareness in the population, although it is likely to underestimate true disease burden, particularly for early or asymptomatic conditions, and should therefore be interpreted with caution (Höhn et al., 2015). As (semi-) automated image-based approaches evolve, they may complement basic self-reported disease assessment and resource and time intensive manual imaging grading for prevalence and incidence estimation by enabling more scalable phenotyping in especially larger datasets. Nonetheless, self-reported prevalence of cataract (4.2%) is consistent with prior evidence indicating that cataract prevalence is often in the single digits in younger populations and increases sharply with age, reaching high levels in older adults . Likewise, the prevalence of self-reported glaucoma (2.0%) aligns with population-based estimates reported in large European cohorts, where glaucoma prevalence generally ranges from approximately 1% to 3%, depending on age and diagnostic definition (Dielemans et al., 1994; Shweikh et al., 2015; Wolfs et al., 2000). The prevalence of self-reported macular degeneration (0.9%) is low compared with image-based estimates in European cohorts, where AMD prevalence increased with age from 3.5% to 17.6% for early AMD and from 0.1% to 9.8% for late AMD, likely because of the comparably young age of the cohort and the occurrence of symptoms in intermediate and late AMD, while patients with early AMD have little to no symptoms.

The ocular data in NAKO also provide important opportunities to develop and validate AI-derived models for automated phenotyping and prediction. The large number of color fundus images from NAKO participants that were classified as gradable by multiple open-source algorithms highlights a key strength of the dataset. This facilitates the development of AI models for eye health research and oculomics approaches. The high quality of the fundus images also made it possible to use deep learning-based tools for automatically extracting morphological features. This allowed us to analyze morphological differences of the retina at a larger scale to answer epidemiological questions, as illustrated for age and sex. Quality assessments by the AI models will be made available via the NAKO Transfer Hub to facilitate further research.

We further demonstrated that the NAKO dataset is suited for developing deep learning techniques to study eye health, including oculomics approaches. Baseline deep learning models trained for predicting age, sex and blood pressure performed similarly to the other approaches based on different fundus image datasets (Ahadi et al., 2023; Poplin et al., 2018; Yu et al., 2025; Zhu et al., 2025). For instance, Poplin et al. (2018) trained an Inception-v3 model on subsets of the UK Biobank (UKB) and the EyePACS dataset (Cuadros & Bresnick, 2009) to predict cardiovascular risk factors, including sex, age and blood pressure. They reported comparable performance to that obtained with the NAKO dataset. They obtained an AUC of 0.97 for sex classification, MAE of 3.26 for age prediction, MAE of 11.35 for systolic blood pressure prediction and MAE of 6.42 for diastolic blood pressure prediction. These values are similar or slightly worse than our best values, with the exception of sex classification. Here, the higher performance of their model was likely due to their use of larger input images with higher resolution. Interestingly, our experiments indicate that a ResNet18 (11.2 M), with a comparably small number of parameters, is sufficient to achieve state-of-the-art results, suggesting larger models could be overparametrized for these specific tasks.

The age-related increase in cup-to-disc ratio and the decline in retinal vessel calibers and AVR observed in this study are consistent with prior population-based and photographic studies describing structural remodeling of the optic nerve head and retina with aging (Garway-Heath et al., 1997; Leung et al., 2003). Sex differences were limited and broadly consistent with previous reports (Leung et al., 2003). These findings should be interpreted in light of the use of measurements of morphological properties obtain using the AutoMorph (Zhou et al., 2022), which, while scalable and reproducible, require further validation against manual expert segmentation, as manual annotation remains the current reference standard. Automatically measured morphological properties will be made available via the NAKO TransferHub as derived variables from the data.

Beyond the baseline characterization presented here, the ophthalmological data availablewithin NAKO will continue to grow, as follow-up examinations are ongoing. In addition, manual image gradings and imaging-derived phenotypes will be released, thereby expanding the scientific scope of the data. Follow-up analyses will permit the assessment of changes over time, and previously developed and adapted AI-based methods will enable the efficient provision of further derived variables for use in scientific analyses and for identifying determinants of progression of ocular disease or maintenance of ocular health at older age.

## Strengths and Limitations

Key strengths of NAKO are its large scale, nationwide coverage, and longitudinal design. Standardized operating procedures across study centers, together with high-quality nonmydriatic color fundus photography and visual acuity assessment, support robust population-based analyses. These data enable the investigation of spatial differences and temporal changes. While this present study focused on automated image analysis pipelines, future releases of manually graded imaging data will improve disease-specific eye phenotyping and validation of imaging-derived endpoints. Recent advances in oculomics and AI highlight the potential of retinal imaging as a biomarker for ocular and systemic diseases. However, its large-scale application in population-based cohorts remains limited. The extensive phenotyping in NAKO will allow for detecting unique, new associations of color fundus image features with a broad set of diseases and health conditions.

Several limitations should be acknowledged. First, retinal imaging within the NAKO ophthalmological assessment is limited to non-mydriatic color fundus photography without complementary modalities, such as optical coherence tomography or wide-field imaging. This restricts detailed structural in depth and peripheral phenotyping. Reliance on self-reported ocular disease may also result in misclassification and under-ascertainment (Foreman et al., 2017; Patty et al., 2012). Second, the width of non-imaging ophthalmic assessment was relatively narrow, encompassing only visual acuity. Third, image quality assessment remains a key unresolved challenge due to the absence of internationally harmonized standards. Although automated quality-control tools can efficiently grade large collections of fundus images and the tools used in this study largely agree on image quality well, remaining inter-algorithm variability raises concerns regarding generalizability and underscores the need for further validation, transparency, and consensus guidance.

## Conclusion and Outlook

The ophthalmological assessment within the NAKO provides a large, standardized, prospective, population-based resource integrating retinal imaging, visual acuity, and comprehensive systemic health measures. We demonstrated its potential as a modern, accessible platform for methodological development and validation in eye health research. Future analyses including follow-up data will provide the opportunity to investigate longitudinal developments among NAKO participants and use the results of the ophthalmological assessment in NAKO for early detection of ocular or systemic diseases.

## Data Availability

All data is accessible to researchers through the official NAKO Transfer Hub (https://transfer.nako.de), subject to application and approval by the NAKO Use and Access Committee, under established governance and data protection frameworks.

https://github.com/berenslab/nako-eye

## Acknowledgements

This project was conducted with data from the German National Cohort (NAKO; www.nako.de; Application No. NAKO-590 and NAKO-810). The NAKO is funded by the Federal Ministry of Research, Technology and Space (BMFTR; project funding reference numbers: 01ER1301A/B/C, 01ER1511D, 01ER1801A/B/C/D and 01ER2301A/B/C), federal states of Germany and the Helmholtz Association, the participating universities and the institutes of the Leibniz Association. We thank all participants who took part in the NAKO study and the staff of this research initiative. The project was additionally funded by the Hertie Foundation. PB is a member of the Excellence Cluster 2064 “Machine Learning – New Perspectives for Science” and the NAKO AI expert group. AS and MU are members of the NAKO retina expert group and the NAKO ophthalmic competence unit. AB is a member of the Digital Clinician Scientist Program at University Medical Center Hamburg-Eppendorf (UKE).

## Contributions

Conceptualization: CR, AB, AS, MU, PB, ML

Methodology: CR, AB, HG, AS, ML, SM

Software: CR, SM

Validation: CR, AB, AS, MU

PB Formal analysis: CR, AB

Investigation: CR, AB, AS

Resources: AS, SM, KB, TB, AE, GG, KHG, CG, KG, AK, TK, JK, ML, CMF, AP, SS, MS, SW, ML, MU, IH

Data Curation: CR, AB, AS, SM, HG

Writing – Original Draft: CR, AB, PB

Writing – Review & Editing: MU, AS, SM, MS, GG, TK, ML, CMF, SS, IH

Visualization: CR

Supervision: MU, PB, GG, CG

Project Administration: AS, MU

Funding acquisition: MU, PB, TK

---

**Supplementary Table 1:** Age distribution of the participants of the baseline from the NAKO Ophthalmology dataset.

| **Age range (years)** | **Male, n (%)** | **Female, n (%)** | **Total, n (%)** |
| --- | --- | --- | --- |
| 10 - 19 | 2 (0.00%) | 2 (0.00%) | 4 (0.01%) |
| 20 - 29 | 2636 (5.44%) | 2343 (4.83%) | 4979 (10.27%) |
| 30 - 39 | 2977 (6.14%) | 2494 (5.15%) | 5471 (11.29%) |
| 40 - 49 | 7136 (14.73%) | 6338 (13.08%) | 13474 (27.80%) |
| 50 - 59 | 6884 (14.21%) | 6394 (13.19%) | 13278 (27.40%) |
| 60 - 69 | 5439 (11.22%) | 4972 (10.26%) | 10411 (21.48%) |
| 70 - 79 | 460 (0.95%) | 383 (0.79%) | 843 (1.74%) |
| All ages | 25534 (52.69%) | 22926 (47.31%) | 48460 (100.00%) |

**Supplementary Table 2:**
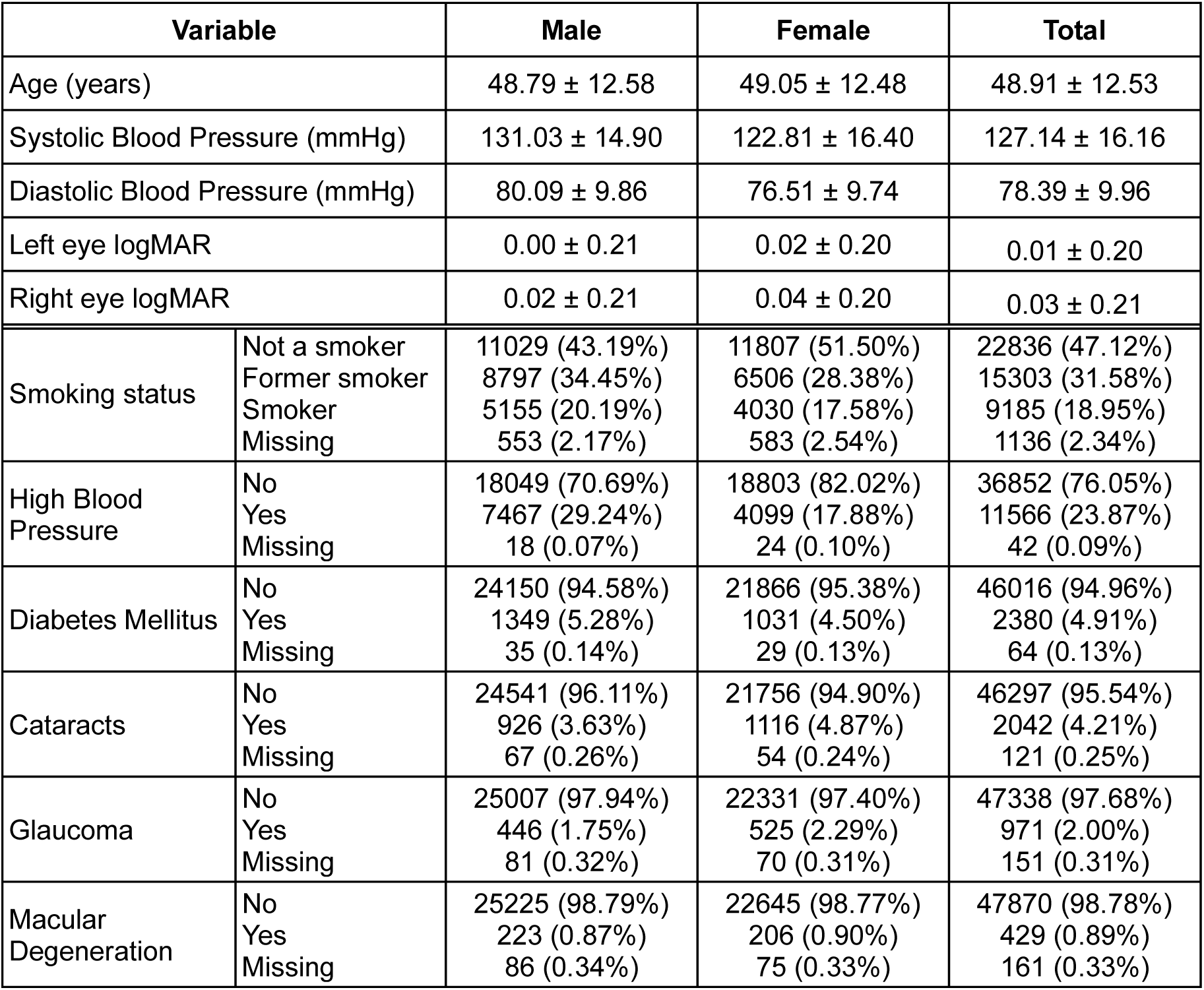
Summary of demographic, systemic, and eye health variables of n=48,460 participants with complete ophthalmological assessment from the baseline in the NAKO Ophthalmology dataset, split by sex. Continuous variables are reported as mean ± sd and discrete variables as n (%). Note: Percentages are relative to the totals in each column.

| Variable |  | Male | Female | Total |
| --- | --- | --- | --- | --- |
| Age (years) | | 48.79 ± 12.58 | 49.05 ± 12.48 | 48.91 ± 12.53 |
| Systolic Blood Pressure (mmHg) | | 131.03 ± 14.90 | 122.81 ± 16.40 | 127.14 ± 16.16 |
| Diastolic Blood Pressure (mmHg) | | 80.09 ± 9.86 | 76.51 ± 9.74 | 78.39 ± 9.96 |
| Left eye logMAR | | 0.00 ± 0.21 | 0.02 ± 0.20 | 0.01 ± 0.20 |
| Right eye logMAR | | 0.02 ± 0.21 | 0.04 ± 0.20 | 0.03 ± 0.21 |
| Smoking status | Not a smoker | 11029 (43.19%) | 11807 (51.50%) | 22836 (47.12%) |
|  | Former smoker | 8797 (34.45%) | 6506 (28.38%) | 15303 (31.58%) |
|  | Smoker | 5155 (20.19%) | 4030 (17.58%) | 9185 (18.95%) |
|  | Missing | 553 (2.17%) | 583 (2.54%) | 1136 (2.34%) |
| High Blood Pressure | No | 18049 (70.69%) | 18803 (82.02%) | 36852 (76.05%) |
|  | Yes | 7467 (29.24%) | 4099 (17.88%) | 11566 (23.87%) |
|  | Missing | 18 (0.07%) | 24 (0.10%) | 42 (0.09%) |
| Diabetes Mellitus | No | 24150 (94.58%) | 21866 (95.38%) | 46016 (94.96%) |
|  | Yes | 1349 (5.28%) | 1031 (4.50%) | 2380 (4.91%) |
|  | Missing | 35 (0.14%) | 29 (0.13%) | 64 (0.13%) |
| Cataracts | No | 24541 (96.11%) | 21756 (94.90%) | 46297 (95.54%) |
|  | Yes | 926 (3.63%) | 1116 (4.87%) | 2042 (4.21%) |
|  | Missing | 67 (0.26%) | 54 (0.24%) | 121 (0.25%) |
| Glaucoma | No | 25007 (97.94%) | 22331 (97.40%) | 47338 (97.68%) |
|  | Yes | 446 (1.75%) | 525 (2.29%) | 971 (2.00%) |
|  | Missing | 81 (0.32%) | 70 (0.31%) | 151 (0.31%) |
| Macular Degeneration | No | 25225 (98.79%) | 22645 (98.77%) | 47870 (98.78%) |
|  | Yes | 223 (0.87%) | 206 (0.90%) | 429 (0.89%) |
|  | Missing | 86 (0.34%) | 75 (0.33%) | 161 (0.33%) |

**Supplementary Table 3:** Summary of demographic, systemic, and eye health variables of n=48,460 participants with complete ophthalmological assessment from the baseline in the NAKO Ophthalmology dataset, split by age group. Continuous variables are reported as mean ± sd and discrete variables as n (%). Note: Percentages are relative to the totals in each column.

| Variable |  | Age range (years) |  |  |  |
| --- | --- | --- | --- | --- | --- |
|  |  | <40 | 40 - 60 | > 60 | All ages |
| Systolic Blood Pressure (mmHg) | | 120.89 ± 12.41 | 126.44 ± 15.47 | 134.6 ± 17.87 | 127.14 ± 16.16 |
| Diastolic Blood Pressure (mmHg) | | 73.99 ± 8.59 | 79.7 ± 9.97 | 79.39 ± 9.98 | 78.39 ± 9.96 |
| Left eye logMAR | | -0.06 ± 0.16 | 0.00 ± 0.20 | 0.09 ± 0.21 | 0.01 ± 0.20 |
| Right eye logMAR | | -0.05 ± 0.17 | 0.02 ± 0.20 | 0.11 ± 0.22 | 0.03 ± 0.21 |
| Sex | Male | 5615 (53.71%) | 14020 (52.41%) | 5899 (52.42%) | 25534 (52.69%) |
|  | Female | 4839 (46.29%) | 12732 (47.59%) | 5355 (47.58%) | 22926 (47.31%) |
| Smoking status | Not a smoker | 5755 (55.05%) | 12286 (45.93%) | 4795 (42.61%) | 22836 (47.12%) |
|  | Former smoker | 2230 (21.33%) | 8524 (31.86%) | 4549 (40.42%) | 15303 (31.58%) |
|  | Smoker | 2338 (22.36%) | 5419 (20.26%) | 1428 (12.69%) | 9185 (18.95%) |
|  | Missing | 131 (1.25%) | 523 (1.95%) | 482 (4.28%) | 1136 (2.34%) |
| High Blood Pressure | No | 9449 (90.39%) | 20472 (76.53%) | 6931 (61.59%) | 36852 (76.05%) |
|  | Yes | 996 (9.53%) | 6257 (23.39%) | 4313 (38.32%) | 11566 (23.87%) |
|  | Missing | 9 (0.09%) | 23 (0.09%) | 10 (0.09%) | 42 (0.09%) |
| Diabetes Mellitus | No | 10297 (98.50%) | 25690 (96.03%) | 10029 (89.11%) | 46016 (94.96%) |
|  | Yes | 149 (1.43%) | 1030 (3.85%) | 1201 (10.67%) | 2380 (4.91%) |
|  | Missing | 8 (0.08%) | 32 (0.12%) | 24 (0.21%) | 64 (0.13%) |
| Cataracts | No | 10428 (99.75%) | 26219 (98.01%) | 9650 (85.75%) | 46297 (95.54%) |
|  | Yes | 17 (0.16%) | 491 (1.84%) | 1534 (13.63%) | 2042 (4.21%) |
|  | Missing | 9 (0.09%) | 42 (0.16%) | 70 (0.62%) | 121 (0.25%) |
| Glaucoma | No | 10408 (99.56%) | 26242 (98.09%) | 10688 (94.97%) | 47338 (97.68%) |
|  | Yes | 29 (0.28%) | 442 (1.65%) | 500 (4.44%) | 971 (2.00%) |
|  | Missing | 17 (0.16%) | 68 (0.25%) | 66 (0.59%) | 151 (0.31%) |
| Macular Degeneration | No | 10423 (99.70%) | 26497 (99.05%) | 10950 (97.30%) | 47870 (98.78%) |
|  | Yes | 16 (0.15%) | 177 (0.66%) | 236 (2.10%) | 429 (0.89%) |
|  | Missing | 15 (0.14%) | 78 (0.29%) | 68 (0.60%) | 161 (0.33%) |

**Supplementary Table 4:** Percentage of good quality images from participants of each age range (n=203,493). Percentages are relative to the total number of available images per age range.

| Age range (years) | FIT | RetiRatey | AutoMorph | VascX | All |
| --- | --- | --- | --- | --- | --- |
| 10 - 19 | 93.8 | 93.8 | 93.8 | 93.8 | 93.8 |
| 20 - 29 | 90.2 | 93.6 | 89.5 | 89.3 | 86.2 |
| 30 - 39 | 86.6 | 91.2 | 86.0 | 85.0 | 81.1 |
| 40 - 49 | 82.6 | 89.2 | 81.5 | 79.3 | 74.6 |
| 50 - 59 | 75.5 | 86.4 | 74.6 | 72.1 | 65.6 |
| 60 - 69 | 60.9 | 80.8 | 61.0 | 60.4 | 50.7 |
| 70 - 79 | 50.8 | 76.9 | 50.5 | 52.1 | 40.8 |

**Supplementary Table 5:** Summary of the GAM fitted to cup-to-disc ratio measurements.

| **A. parametric coefficients** | **Estimate** | **Std. Error** | **t-value** | **p-value** |
| --- | --- | --- | --- | --- |
| (Intercept) | 0.50 | 0.00 | 909.19 | < 0.0001 |
| sex Male | 0.00 | 0.00 | 4.65 | < 0.0001 |
| **B. smooth terms** | **edf** | **Ref.df** | **F-value** | **p-value** |
| s(age): sex Female | 1.01 | 1.02 | 197.52 | < 0.0001 |
| s(age): sex Male | 2.82 | 3.52 | 50.89 | < 0.0001 |
| **C. other stats** | **R2** | **Deviance explained** | **REML** | **n** |
|  | 0.00925 | 0.94% | -36976 | 28220 |

**Supplementary Table 6:** Summary of the GAM fitted to central retinal artery equivalent (CRAE) measurements.

| **A. parametric coefficients** | **Estimate** | **Std. Error** | **t-value** | **p-value** |
| --- | --- | --- | --- | --- |
| (Intercept) | 155.35 | 0.15 | 1071.63 | < 0.0001 |
| sex Male | -1.89 | 0.20 | -9.27 | < 0.0001 |
| **B. smooth terms** | **edf** | **Ref.df** | **F-value** | **p-value** |
| s(age): sex Female | 4.88 | 5.93 | 1323.27 | < 0.0001 |
| s(age): sex Male | 4.32 | 5.31 | 1794.85 | < 0.0001 |
| **C. other stats** | **R2** | **Deviance explained** | **REML** | **n** |
|  | 0.104 | 10.50% | 115850 | 27297 |

**Supplementary Table 7:** Summary of the GAM fitted to arteriolar-to-venular ratio measurements.

| **A. parametric coefficients** | **Estimate** | **Std. Error** | **t-value** | **p-value** |
| --- | --- | --- | --- | --- |
| (Intercept) | 0.68 | 0.00 | 1003.38 | < 0.0001 |
| sex Male | -0.01 | 0.00 | -10.53 | < 0.0001 |
| **B. smooth terms** | **edf** | **Ref.df** | **F-value** | **p-value** |
| s(age): sex Female | 3.81 | 4.72 | 238.12 | < 0.0001 |
| s(age): sex Male | 4.15 | 5.12 | 442.26 | < 0.0001 |
| **C. other stats** | **R2** | **Deviance explained** | **REML** | **n** |
|  | 0.0279 | 2.82% | -30337 | 27114 |

**Supplementary Table 8:** Summary of the GAM fitted to vessel density measurements.

| **A. parametric coefficients** | **Estimate** | **Std. Error** | **t-value** | **p-value** |
| --- | --- | --- | --- | --- |
| (Intercept) | 0.01 | 0.00 | 638.68 | < 0.0001 |
| sex Male | 0.00 | 0.00 | -3.54 | 0.0004 |
| **B. smooth terms** | **edf** | **Ref.df** | **F-value** | **p-value** |
| s(age): sex Female | 4.39 | 5.39 | 571.31 | < 0.0001 |
| s(age): sex Male | 3.71 | 4.61 | 613.21 | < 0.0001 |
| **C. other stats** | **R2** | **Deviance explained** | **REML** | **n** |
|  | 0.0399 | 4.02% | -149620 | 28608 |

## Literature

Ahadi, S., Wilson, K. A., Babenko, B., McLean, C. Y., Bryant, D., Pritchard, O., Kumar, A., Carrera, E. M., Lamy, R., Stewart, J. M., Varadarajan, A., Berndl, M., Kapahi, P., & Bashir, A. (2023). Longitudinal fundus imaging and its genome-wide association analysis provide evidence for a human retinal aging clock. eLife, 12, e82364. 10.7554/eLife.82364

Beuse, A., Hoppert, I., Wolfram, C., Spitzer, M. S., Birtel, J., & Grohmann, C. (2025). Hamburg City Health Study: Design and ocular baseline characteristics of the first 10,000 participants. Investigative Ophthalmology & Visual Science, 66(8), 2009–2009.

Brandl, C., Brücklmayer, C., Günther, F., Zimmermann, M. E., Küchenhoff, H., Helbig, H., Weber, B. H. F., Heid, I. M., & Stark, K. J. (2019). Retinal Layer Thicknesses in Early Age-Related Macular Degeneration: Results From the German AugUR Study. Investigative Ophthalmology & Visual Science, 60(5), 1581–1594. 10.1167/iovs.18-25332

Chen, Z., Chen, J., Collins, R., Guo, Y., Peto, R., Wu, F., Li, L., & on behalf of the China Kadoorie Biobank (CKB) collaborative group. (2011). China Kadoorie Biobank of 0.5 million people: Survey methods, baseline characteristics and long-term follow-up. International Journal of Epidemiology, 40(6), 1652–1666. 10.1093/ije/dyr120

Chollet, F. (2017). Xception: Deep Learning With Depthwise Separable Convolutions. 1251–1258. https://openaccess.thecvf.com/content_cvpr_2017/html/Chollet_Xception_Deep_Learning_CVPR_2017_paper.html

Chua, S. Y. L., Thomas, D., Allen, N., Lotery, A., Desai, P., Patel, P., Muthy, Z., Sudlow, C., Peto, T., Khaw, P. T., & Foster, P. J. (2019). Cohort profile: Design and methods in the eye and vision consortium of UK Biobank. BMJ Open, 9(2), e025077. 10.1136/bmjopen-2018-025077

Cuadros, J., & Bresnick, G. (2009). EyePACS: An Adaptable Telemedicine System for Diabetic Retinopathy Screening. Journal of Diabetes Science and Technology, 3(3), 509–516. 10.1177/193229680900300315

Dielemans, I., Vingerling, J. R., Wolfs, R. C. W., Hofman, A., Grobbee, D. E., & de Jong, P. T. V. M. (1994). The Prevalence of Primary Open-angle Glaucoma in a Population-based Study in The Netherlands: The Rotterdam Study. Ophthalmology, 101(11), 1851–1855. 10.1016/S0161-6420(94)31090-6

Dosovitskiy, A., Beyer, L., Kolesnikov, A., Weissenborn, D., Zhai, X., Unterthiner, T., Dehghani, M., Minderer, M., Heigold, G., Gelly, S., Uszkoreit, J., & Houlsby, N. (2020, October 2). An Image is Worth 16×16 Words: Transformers for Image Recognition at Scale. International Conference on Learning Representations. https://openreview.net/forum?id=YicbFdNTTy

Foreman, J., Xie, J., Keel, S., van Wijngaarden, P., Taylor, H. R., & Dirani, M. (2017). The validity of self-report of eye diseases in participants with vision loss in the National Eye Health Survey. Scientific Reports, 7(1), 8757. 10.1038/s41598-017-09421-9

Fu, H., Wang, B., Shen, J., Cui, S., Xu, Y., Liu, J., & Shao, L. (2019). Evaluation of Retinal Image Quality Assessment Networks in Different Color-Spaces. In D. Shen, T. Liu, T. M. Peters, L. H. Staib, C. Essert, S. Zhou, P.-T. Yap, & A. Khan (Eds.), Medical Image Computing and Computer Assisted Intervention – MICCAI 2019 (pp. 48–56). Springer International Publishing. 10.1007/978-3-030-32239-7_6

Garway-Heath, D. F., Wollstein, G., & Hitchings, R. A. (1997). Aging changes of the optic nerve head in relation to open angle glaucoma. British Journal of Ophthalmology, 81(10), 840–845. 10.1136/bjo.81.10.840

Gervelmeyer, J., Müller, S., Huang, Z., & Berens, P. (2025). Fundus Image Toolbox: A Python package for fundus image processing. Journal of Open Source Software, 10(108), 7101. 10.21105/joss.07101

He, K., Zhang, X., Ren, S., & Sun, J. (2016). Deep Residual Learning for Image Recognition. 770–778. https://openaccess.thecvf.com/content_cvpr_2016/html/He_Deep_Residual_Learning_CVPR_2016_paper.html

Höhn, R., Kottler, U., Peto, T., Blettner, M., Münzel, T., Blankenberg, S., Lackner, K. J., Beutel, M., Wild, P. S., & Pfeiffer, N. (2015). The Ophthalmic Branch of the Gutenberg Health Study: Study Design, Cohort Profile and Self-Reported Diseases. PLOS ONE, 10(3), e0120476. 10.1371/journal.pone.0120476

Holle, R., Happich, M., Löwel, H., Wichmann, H. E., & Group, for the M. S. (2005). KORA - A Research Platform for Population Based Health Research. Das Gesundheitswesen, 67(S 01), 19–25. 10.1055/s-2005-858235

Jagodzinski, A., Johansen, C., Koch-Gromus, U., Aarabi, G., Adam, G., Anders, S., Augustin, M., der Kellen, R. B., Beikler, T., Behrendt, C.-A., Betz, C. S., Bokemeyer, C., Borof, K., Briken, P., Busch, C.-J., Büchel, C., Brassen, S., Debus, E. S., Eggers, L., … Blankenberg, S. (2020). Rationale and Design of the Hamburg City Health Study. European Journal of Epidemiology, 35(2), 169–181. 10.1007/s10654-019-00577-4

Knudtson, M. D., Lee, K. E., Hubbard, L. D., Wong, T. Y., Klein, R., & Klein, B. E. K. (2003). Revised formulas for summarizing retinal vessel diameters. Current Eye Research, 27(3), 143–149. 10.1076/ceyr.27.3.143.16049

Leitritz, M. A., Hense, H.-W., Schiefer, U., Nagel, M., Greiser, H., Linseisen, J., Heid, I., Fischer, B., Thierry, S., Bartz-Schmidt, K. U., & Ueffing, M. (2013a). [Development and first results of fast and cost-effective examination methods for an ophthalmological screening within the National Cohort]. Klinische Monatsblatter fur Augenheilkunde, 230(12), 1238–1246. 10.1055/s-0033-1350685

Leitritz, M. A., Hense, H.-W., Schiefer, U., Nagel, M., Greiser, H., Linseisen, J., Heid, I., Fischer, B., Thierry, S., Bartz-Schmidt, K. U., & Ueffing, M. (2013b). Entwicklung und erste Anwendungserfahrungen einer schnellen und kostengünstigen ophthalmologischen Untersuchung für die Nationale Kohorte. Klinische Monatsblätter für Augenheilkunde, 230(12), 1238–1246. 10.1055/s-0033-1350685

Leung, H., Wang, J. J., Rochtchina, E., Tan, A. G., Wong, T. Y., Klein, R., Hubbard, L. D., & Mitchell, P. (2003). Relationships between Age, Blood Pressure, and Retinal Vessel Diameters in an Older Population. Investigative Ophthalmology & Visual Science, 44(7), 2900–2904. 10.1167/iovs.02-1114

Liu, R., Wang, X., Wu, Q., Dai, L., Fang, X., Yan, T., Son, J., Tang, S., Li, J., Gao, Z., Galdran, A., Poorneshwaran, J. M., Liu, H., Wang, J., Chen, Y., Porwal, P., Wei Tan, G. S., Yang, X., Dai, C., … Zhang, P. (2022). DeepDRiD: Diabetic Retinopathy—Grading and Image Quality Estimation Challenge. Patterns, 3(6), 100512. 10.1016/j.patter.2022.100512

Liu, Z., Lin, Y., Cao, Y., Hu, H., Wei, Y., Zhang, Z., Lin, S., & Guo, B. (2021). Swin Transformer: Hierarchical Vision Transformer Using Shifted Windows. 10012–10022. https://openaccess.thecvf.com/content/ICCV2021/html/Liu_Swin_Transformer_Hierarchical_Vision_Transformer_Using_Shifted_Windows_ICCV_2021_paper.html

Loeffler, M., Engel, C., Ahnert, P., Alfermann, D., Arelin, K., Baber, R., Beutner, F., Binder, H., Brähler, E., Burkhardt, R., Ceglarek, U., Enzenbach, C., Fuchs, M., Glaesmer, H., Girlich, F., Hagendorff, A., Häntzsch, M., Hegerl, U., Henger, S., … Thiery, J. (2015). The LIFE-Adult-Study: Objectives and design of a population-based cohort study with 10,000 deeply phenotyped adults in Germany. BMC Public Health, 15(1), 691. 10.1186/s12889-015-1983-z

Mauschitz, M. M., Lohner, V., Koch, A., Stöcker, T., Reuter, M., Holz, F. G., Finger, R. P., & Breteler, M. M. B. (2022). Retinal layer assessments as potential biomarkers for brain atrophy in the Rhineland Study. Scientific Reports, 12(1), 2757. 10.1038/s41598-022-06821-4

Oraá, R. R., García, M., Oraá-Pérez, J., López, M. I., & Hornero, R. (2020). Automatic fundus image quality assessment: Diagnostic accuracy in clinical practice. Investigative Ophthalmology & Visual Science, 61(7), 2033–2033.

Patty, L., Wu, C., Torres, M., Azen, S., & Varma, R. (2012). Validity of Self-reported Eye Disease and Treatment in a Population-based Study: The Los Angeles Latino Eye Study. Ophthalmology, 119(9), 1725–1730. 10.1016/j.ophtha.2012.02.029

Peters, A., Peters, A., Greiser, K. H., Göttlicher, S., Ahrens, W., Albrecht, M., Bamberg, F., Bärnighausen, T., Becher, H., Berger, K., Beule, A., Boeing, H., Bohn, B., Bohnert, K., Braun, B., Brenner, H., Bülow, R., Castell, S., Damms-Machado, A., … German National Cohort (NAKO) Consortium. (2022). Framework and baseline examination of the German National Cohort (NAKO). European Journal of Epidemiology, 37(10), 1107–1124. 10.1007/s10654-022-00890-5

Poplin, R., Varadarajan, A. V., Blumer, K., Liu, Y., McConnell, M. V., Corrado, G. S., Peng, L., & Webster, D. R. (2018). Prediction of cardiovascular risk factors from retinal fundus photographs via deep learning. Nature Biomedical Engineering, 2(3), Article 3. 10.1038/s41551-018-0195-0

Rach, S., Sand, M., Reineke, A., Becher, H., Greiser, K. H., Wolf, K., Wirkner, K., Schmidt, C. O., Schipf, S., Jöckel, K.-H., Krist, L., Ahrens, W., Brenner, H., Castell, S., Gastell, S., Harth, V., Holleczek, B., Ittermann, T., Janisch-Fabian, S., … Günther, K. (2025). The baseline examinations of the German National Cohort (NAKO): Recruitment protocol, response, and weighting. European Journal of Epidemiology, 40(4), 475–489. 10.1007/s10654-025-01219-8

Schipf, S., Schöne, G., Schmidt, B., Günther, K., Stübs, G., Greiser, K. H., Bamberg, F., Meinke-Franze, C., Becher, H., Berger, K., Brenner, H., Castell, S., Damms-Machado, A., Fischer, B., Franzke, C.-W., Fricke, J., Gastell, S., Günther, M., Hoffmann, W., … Ahrens, W. (2020). Die Basiserhebung der NAKO Gesundheitsstudie: Teilnahme an den Untersuchungsmodulen, Qualitätssicherung und Nutzung von Sekundärdaten. Bundesgesundheitsblatt - Gesundheitsforschung - Gesundheitsschutz, 63(3), 254–266. 10.1007/s00103-020-03093-z

Shweikh, Y., Ko, F., Chan, M. P. Y., Patel, P. J., Muthy, Z., Khaw, P. T., Yip, J., Strouthidis, N., & Foster, P. J. (2015). Measures of socioeconomic status and self-reported glaucoma in the UK Biobank cohort. Eye, 29(10), 1360–1367. 10.1038/eye.2015.157

Stark, K., Olden, M., Brandl, C., Dietl, A., Zimmermann, M. E., Schelter, S. C., Loss, J., Leitzmann, M. F., Böger, C. A., Luchner, A., Kronenberg, F., Helbig, H., Weber, B. H. F., & Heid, I. M. (2015). The German AugUR study: Study protocol of a prospective study to investigate chronic diseases in the elderly. BMC Geriatrics, 15(1), 130. 10.1186/s12877-015-0122-0

Stein, M. J., Fischer, B., Bohmann, P., Ahrens, W., Berger, K., Brenner, H., Günther, K., Harth, V., Heise, J.-K., Karch, A., Klett-Tammen, C. J., Koch-Gallenkamp, L., Krist, L., Lieb, W., Meinke-Franze, C., Michels, K. B., Mikolajczyk, R., Nimptsch, K., Obi, N., … Sedlmeier, A. M. (2024). Differences in Anthropometric Measures Based on Sex, Age, and Health Status. Deutsches Ärzteblatt International, 121(7), 207–213. 10.3238/arztebl.m2024.0016

Sudlow, C., Gallacher, J., Allen, N., Beral, V., Burton, P., Danesh, J., Downey, P., Elliott, P., Green, J., Landray, M., Liu, B., Matthews, P., Ong, G., Pell, J., Silman, A., Young, A., Sprosen, T., Peakman, T., & Collins, R. (2015). UK Biobank: An Open Access Resource for Identifying the Causes of a Wide Range of Complex Diseases of Middle and Old Age. PLOS Medicine, 12(3), e1001779. 10.1371/journal.pmed.1001779

Szegedy, C., Vanhoucke, V., Ioffe, S., Shlens, J., & Wojna, Z. (2016). Rethinking the Inception Architecture for Computer Vision. 2818–2826. https://www.cv-foundation.org/openaccess/content_cvpr_2016/html/Szegedy_Rethinking_the_Inception_CVPR_2016_paper.html

Tan, M., & Le, Q. (2021). EfficientNetV2: Smaller Models and Faster Training. Proceedings of the 38th International Conference on Machine Learning, 10096–10106. https://proceedings.mlr.press/v139/tan21a.html

The National German Cohort Consortium. (2014). The German National Cohort: Aims, study design and organization. European Journal of Epidemiology, 29(5), 371–382. 10.1007/s10654-014-9890-7

Vargas Quiros, J., Liefers, B., van Garderen, K. A., Vermeulen, J. P., & Klaver, C. (2025). VascX Models: Deep Ensembles for Retinal Vascular Analysis From Color Fundus Images. Translational Vision Science & Technology, 14(7), 19. 10.1167/tvst.14.7.19

Warwick, A. N., Curran, K., Hamill, B., Stuart, K., Khawaja, A. P., Foster, P. J., Lotery, A. J., Quinn, M., Madhusudhan, S., Balaskas, K., & Peto, T. (2023). UK Biobank retinal imaging grading: Methodology, baseline characteristics and findings for common ocular diseases. Eye, 37(10), 2109–2116. 10.1038/s41433-022-02298-7

Wild, P. S., Zeller, T., Beutel, M., Blettner, M., Dugi, K. A., Lackner, K. J., Pfeiffer, N., Münzel, T., & Blankenberg, S. (2012). [The Gutenberg Health Study]. Bundesgesundheitsblatt, Gesundheitsforschung, Gesundheitsschutz, 55(6–7), 824–829. 10.1007/s00103-012-1502-7

Wolfs, R. C. W., Borger, P. H., Ramrattan, R. S., Klaver, C. C. W., Hulsman, C. A. A., Hofman, A., Vingerling, J. R., Hitchings, R. A., & de Jong, P. T. V. M. (2000). Changing Views on Open-Angle Glaucoma: Definitions and Prevalences—The Rotterdam Study. Investigative Ophthalmology & Visual Science, 41(11), 3309–3321.

Wood, S. N. (2017). Generalized Additive Models: An Introduction with R*, Second Edition* (2nd ed.). Chapman and Hall/CRC. 10.1201/9781315370279

Yu, Z., Chen, R., Gui, P., Wang, W., Razzak, I., Alinejad-Rokny, H., Zeng, X., Shang, X., Zhang, L., Yang, X., Yu, H., Huang, W., Lu, H., van Wijngaarden, P., He, M., Zhu, Z., & Ge, Z. (2025). A cross population study of retinal aging biomarkers with longitudinal pre-training and label distribution learning. Npj Digital Medicine, 8(1), 344. 10.1038/s41746-025-01751-7

Zhou, Y., Wagner, S. K., Chia, M. A., Zhao, A., Woodward-Court, P., Xu, M., Struyven, R., Alexander, D. C., & Keane, P. A. (2022). AutoMorph: Automated Retinal Vascular Morphology Quantification Via a Deep Learning Pipeline. Translational Vision Science & Technology, 11(7), 12. 10.1167/tvst.11.7.12

Zhu, Z., Wang, Y., Qi, Z., Hu, W., Zhang, X., Wagner, S. K., Wang, Y., Ran, A. R., Ong, J., Waisberg, E., Masalkhi, M., Suh, A., Tham, Y. C., Cheung, C. Y., Yang, X., Yu, H., Ge, Z., Wang, W., Sheng, B., … Wong, T. Y. (2025). Oculomics: Current concepts and evidence. Progress in Retinal and Eye Research, 106, 101350. 10.1016/j.preteyeres.2025.101350

